# Depressive and anxiety symptoms during the COVID-19 pandemic: A two-year follow-up

**DOI:** 10.1101/2022.05.24.22275529

**Authors:** Feifei Bu, Andrew Steptoe, Daisy Fancourt

## Abstract

**Background:** There has been much research into the mental health impact of the COVID-19 pandemic and how it is related to time-invariant individual characteristics (e.g. age and gender). However, there is still a lack of research showing long-term trajectories of mental health across different stages of the pandemic. And little is known regarding the longitudinal association of time-varying contextual and individual factors (e.g. COVID-19 policy response and pandemic intensity) with mental health outcomes. This study aimed to provide a longitudinal profile of how depressive and anxiety symptoms changed by month between March 2020 and April 2022, and to examine their longitudinal associations with time-varying contextual and individual level factors.

**Methods and findings:** Drawing data from a large panel study of over 58,000 adults living in England, we showed that mental health changes were largely in line with changes in COVID-19 policy response and pandemic intensity. Further, data were analysed using fixed-effects, with models fitted separately across three stages of the COVID-19 pandemic. We found that more stringent policy response was associated with increased mental health symptoms, in particular during lockdown periods. Higher COVID-19 deaths were also associated with poorer mental health, but this association weakened over time. Finally, there was also evidence for the longitudinal association of mental health with individual level factors, including confidence in government/healthcare/essentials, COVID-19 knowledge, COVID-19 stress, COVID-19 infection and social support.

**Conclusions:** Our results provided empirical evidence on how changes in contextual and individual level factors were related to depressive and anxiety symptoms. While some factors clearly acted as consistent predictors of mental health during a pandemic, other factors were dependent on the specific situations occurring within society. This could provide important implications for policy making and for a better understanding of mental health of the general public during a national or global health crisis.

## Introduction

The COVID-19 pandemic has caused unprecedented disruptions to the global economy and people’s everyday lives. It has presented various psychological challenges, such as uncertainty and fear about the virus, enforced social isolation caused by lockdown and social distancing measures, financial adversities, reduced access to healthcare services and so forth [1, 2]. At the start of the pandemic, this led to wide-spread concerns about what the global mental health impact would be [3, 4], with subsequent empirical evidence showing worsening mental health during, compared to prior to, the pandemic [5–7]. However, in the two years since, the political and public health context has changed substantially with the roll out of vaccinations and loosening of social restrictions in many countries, despite increasing case and death rates. Over this period, there has been some evidence from longitudinal studies for a gradual improvement in mental health from the start of the pandemic [2, 8, 9]. But the follow-up periods of such studies have been relatively short to provide a long-term perspective of mental health changes across different stages of the COVID-19 pandemic. To the best of our knowledge, no study to date has shown detailed longitudinal changes in public mental health over the first two years of the pandemic.

Further, numerous studies have shown how mental health during the pandemic has been related sociodemographic factors (e.g. age, gender, socioeconomic position, health conditions etc.) [2, 7], most of which are time-invariant, focusing on individual characteristics. But there has been a much smaller literature on the relationship of time-varying and, in particular, contextual factors with mental health, such as COVID-19 cases, deaths and policy responses. A meta-analysis of depression outcomes across 33 countries found that the prevalence of clinical depression was significantly lower in countries where stringent policies were implemented in early 2020 [10]. This is consistent with another cross-country study showing that overall stringency was associated with lower Google searches for ‘depression’, but no evidence for ‘anxiety’ in 2020 [11]. However, a recent study using longitudinal data between April 2020 and June 2021 from 15 countries showed that higher policy stringency and number of deaths were both associated with poor mental health outcomes [12]. These inconsistencies could reflect the differential association of mental health with contextual factors across difference stages of the COVID-19 pandemic, and potential differences between depression and anxiety. Further, it is relevant to consider whether predictors of mental health during the pandemic have changed as the context of COVID-19 has shifted. Indeed, research into behaviours during the pandemic (e.g. compliance with rules) has shown that predictors have not been constant but rather have been context-specific [13]. This effect may also be salient for psychological experiences and has important consequences for pandemic planning.

Therefore, drawing data from a large panel study of over 58,000 adults with a follow-up of 25 months, this study first aimed to show how depressive and anxiety symptoms changed by month between March 2020 and April 2022 in England. Second, it aimed to examine how changes in depressive and anxiety symptoms were associated with changes in time-varying contextual and individual level factors. Moreover, we were interested to assess if any of these longitudinal associations differed across different stages of the pandemic. For this, fixed-effects models were fitted across three distinct periods between March 2020 and November 2021.

## Method

### Data

This study analysed data from the University College London (UCL) COVID-19 Social Study (CSS), a large panel study of the psychological and social experiences of over 75,000 adults (aged 18+) in the UK during the COVID-19 pandemic. The study commenced on 21 March 2020 and involved weekly online data collection until August 2020 and then monthly (four-weekly) until November 2021. When the survey was converted to monthly, participants were randomly assigned into four groups. Each received the survey invitation and reminder at a different week of the month to have survey responses spread out over different days (averaging 970 participants per day during monthly data collection compared to 4300 per day during weekly data collection). Following the monthly data collection, additional follow-ups were carried out in January 2022 and March 2022. The study did not use a random sample design and therefore the original sample is not representative of the UK population. However, the study had a three-fold recruitment strategy that was designed to maximise the heterogeneity of the sample and ensure representation from vulnerable and marginalised groups, and all data analysed from the study are weighted to match population proportions. First, convenience sampling was used, including promoting the study through existing networks and mailing lists (including large databases of adults who had previously consented to be involved in health research across the UK), print and digital media coverage, and social media. Second, more targeted recruitment was undertaken focusing on (i) individuals from a low-income background, (ii) individuals with no or few educational qualifications, and (iii) individuals who were unemployed. Third, the study was promoted via partnerships with third sector organisations to vulnerable groups, including adults with pre-existing mental health conditions, older adults, carers, and people experiencing domestic violence or abuse. The study was approved by the UCL Research Ethics Committee [12467/005] and all participants gave informed consent. A full protocol for the study is available online at https://osf.io/jm8ra/.

This study focused on participants participating in the study who were living in England (N=59,810). After excluding participants missing data in depressive or anxiety symptoms or any of variables included in weighting (see Statistical Analysis below), we had an analytical sample of 58,442 unique participants and 957,110 total observations.

### Measures

#### Mental health outcomes

Depressive symptoms were measured using the Patient Health Questionnaire (PHQ-9) [14]; a standardised instrument for screening for depression in primary care. Unlike the original PHQ-9, the current study enquired about symptoms ‘over the last week’ instead of ‘over the last two weeks’ as data were initially collected weekly. The questionnaire includes 9 items with 4-point responses ranging from ‘not at all’ to ‘nearly every day’. Higher overall scores indicate more depressive symptoms, ranging from 0 to 27.

Anxiety symptoms were measured using the Generalized Anxiety Disorder assessment (GAD-7) [15]; a well-validated tool used to screen for generalized anxiety disorder in clinical practice and research. These questions were also worded as ‘over the last week’ for the same reason as the depression items. The GAD-7 comprises 7 items with 4-point responses ranging from ‘not at all’ to ‘nearly every day’, with higher overall scores indicating more symptoms of anxiety, ranging from 0 to 21.

#### Contextual predictors

##### Policy responses

Stringency Index was obtained from the Oxford COVID-19 Government Response Tracker (OxCGRT) [16]. This records the strictness of governmental policies that primarily restrict individual’s behaviours through a composite measure of nine response metrics, such as stay-at-home orders, workplace closure, cancellation of public events, public information campaigns, and so forth. It ranges from 0 to 100, with a higher value indicating a stricter response. The index is available at sub-national level in the UK. This study used data exclusively from England.

Vaccination was defined as the cumulative number of vaccines given including all doses by published date (in England) obtained from the UK government website for data on COVID [17].

##### Pandemic intensity

Daily Covid-19 cases was defined as number of new cases per day based on data published date. Daily Covid-19 deaths was defined as new deaths within 28 days of positive test. Both measures (in England) were obtained from the UK government website [17]. All contextual variables are available on daily basis which were linked to the individual data from CSS based on survey dates.

#### Individual predictors

At an individual level, we measured people’s confidence in the government, healthcare services and access to essentials (e.g. access to food, water, medicines, deliveries). Participants were asked to rate their confidence levels for each on a scale to 1 (not at all confident) to 7 (very confident). Participants were also asked to rate their knowledge level on COVID-19 on a scale of 1 to 7. For COVID-19 stress, people were asked if they had been worrying about catching COVID-19 (0=no, 1=yes, minor stress, 2=yes, major stress) or becoming seriously ill from COVID-19 (0=no, 1=yes, minor stress, 2=yes, major stress). These were combined into a total stress score ranging from 0 to 4. Moreover, we included self-reported COVID-19 infection. During the weekly data collection (March-August 2020), participants were asked if they had had COVID-19; whereas during the four-weekly phrase, they were asked if they had had COVID-19 in the past month. This was coded as a binary variable (0=no/not know of, 1=diagnosed/not formally diagnosed but suspected). Finally, we considered perceived social support as a mental health predictor. This was measured by an adapted version of the six-item short form of Perceived Social Support Questionnaire, with a high reliability (α=0.90) [18] Each item was rated on a five-point scale from ‘not true at all’ to ‘very true’. A total score was generated based on confirmatory factor analysis, with higher scores indicating higher levels of support. All predictors (except for COVID-19 infection) were standardised to have a mean of 0 and standard deviation of 1. Full questions and responses for each item described above are available online at https://osf.io/jm8ra/.

### Statistical Analysis

We started with descriptive analyses showing mental health trends over time. Data were analysed in four-week periods from March 2020 to November 2021, with additional follow-ups in January and March 2022. Weekly data between March-August 2020 were aggregated by taking means within each individual and each wave of data were weighted separately to the proportions of gender, age, ethnicity and education in the English adult population [19].

Next, data were analysed using fixed-effects (FE) models. FE analysis uses only within-individual variation, which automatically controlling for observed or unobserved individual heterogeneities [20]. It addresses how the change in a predictor is related to the change in mental health outcomes. Further, we were interested in if any longitudinal association differed across time. Therefore, FE models were fitted separately across three periods based on study design and changes in policy responses (see Figure S1 in the Supplement): 1) 1^st^ national lockdown (until the end of weekly data collection): 21/03/2020-23/08/2020 (weekly data, N=45838, mean number of time points (T)=11.5), 2) 2^nd^ and 3^rd^ national lockdown: 21/09/2020-11/04/2021 (four-weekly data, N=26175, T=6.1), 3) ‘freedom’ period: 12/04/2021-14/11/2021 (four-weekly data, N=21194, T=6.3). In each dataset for the FE analyses, we had further excluded participants with missing values in any of the predictors and those less than two time points. These three datasets were weighted separately. To address multiplicity, we provided adjusted p values (q values) controlling for the positive false discovery rate, using the Benjamini-Yekutieli method implemented with the ‘qqvalue’ command [21]. All analyses were conducted using Stata V17.

## Results

### Descriptive statistics

Baseline characteristics of participants of different study periods are shown in Table S1 in the Supplement. After weighting, the three cohorts showed sociodemographic profiles in line with national statistics. These and other time-invariant measures (observed or unobserved) were controlled for but not estimated in FE analyses. For time-varying measures, the within and between person variations are reported in Table 1. The mean depressive symptoms were slightly higher during the two lockdown periods 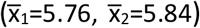 compared to the ‘freedom’ period 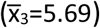. In all periods, the variation in depressive symptoms was mostly related to the differences between participants (ρ=0.82). Similarly, anxiety symptoms also appeared to be higher during lockdowns, with variation mostly due to between person differences. As expected, means of the contextual predictors varied substantially across three study periods. For example, national policies were the strictest during the 2^nd^ and 3^rd^ lockdowns period 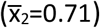 and least restricted in the ‘freedom’ period 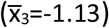. Daily COVID-19 cases were low during the 1^st^ lockdown 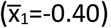, but much higher later on 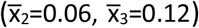. Daily deaths due to COVID-19 were higher during lockdown periods 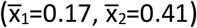, but much lower in the last study period 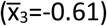. This could be partially related to vaccination, which started on 8 December 2020. Therefore, vaccination was invariant in Period I and much lower in Period II than Period III. The variation in the contextual predictors was mostly related to differences within participants over time. For individual level predictors, changes over time were also observed. However, variation in most of these variables were largely due to between participant differences.

**Table 1.** Descriptive statistics of time-varying variables across three periods (weighted)

|  | Period I: 1<sup>st</sup> lockdown<br>(21/03/2020-23/08/2020)<br>(N=45 838, T=11.5) |  |  |  | Period II: 2<sup>nd</sup> & 3<sup>rd</sup> lockdowns<br>(21/09/2020-11/04/2021)<br>(N=26 175, T=6.1) |  |  |  | Period III: freedom<br>(12/04/2021-14/11/2021)<br>(N=21 194, T=6.3) |  |  |  |
| --- | --- | --- | --- | --- | --- | --- | --- | --- | --- | --- | --- | --- |
| | Mean<br>$\bar{X}_1$ | Between<br>$\sigma_{u1}$ | Within<br>$\sigma_{e1}$ | Ratio<br>$\rho_1$ | Mean<br>$\bar{X}_2$ | Between<br>$\sigma_{u2}$ | Within<br>$\sigma_{e2}$ | Ratio<br>$\rho_2$ | Mean<br>$\bar{X}_3$ | Between<br>$\sigma_{u3}$ | Within<br>$\sigma_{e3}$ | Ratio<br>$\rho_3$ |
| Depressive symptoms | 5.76 | 5.36 | 2.50 | 0.82 | 5.84 | 5.18 | 2.44 | 0.82 | 5.69 | 5.06 | 2.37 | 0.82 |
| Anxiety symptoms | 4.29 | 4.77 | 2.22 | 0.82 | 4.40 | 4.54 | 2.18 | 0.81 | 4.10 | 4.38 | 2.10 | 0.81 |
| Stringency index (std) | 0.35 | 0.32 | 0.44 | 0.34 | 0.71 | 0.15 | 0.51 | 0.08 | -1.13 | 0.41 | 1.00 | 0.14 |
| Vaccination (std) | -0.55 | 0.00 | 0.00 | -- | -0.34 | 0.12 | 0.31 | 0.12 | 1.30 | 0.19 | 0.45 | 0.16 |
| New cases per day (std) | -0.40 | 0.03 | 0.03 | 0.39 | 0.06 | 0.11 | 0.42 | 0.07 | 0.12 | 0.17 | 0.42 | 0.14 |
| New deaths per day (std) | 0.17 | 0.69 | 0.96 | 0.34 | 0.41 | 0.45 | 1.28 | 0.11 | -0.61 | 0.07 | 0.18 | 0.14 |
| Confidence: government (std) | 0.18 | 0.87 | 0.48 | 0.77 | -0.06 | 0.91 | 0.41 | 0.83 | 0.09 | 0.96 | 0.38 | 0.87 |
| Confidence: healthcare (std) | 0.12 | 0.85 | 0.55 | 0.70 | -0.02 | 0.87 | 0.57 | 0.70 | 0.14 | 0.88 | 0.51 | 0.75 |
| Confidence: essential (std) | 0.06 | 0.84 | 0.58 | 0.68 | 0.10 | 0.85 | 0.54 | 0.71 | -0.00 | 0.86 | 0.70 | 0.60 |
| COVID-19 knowledge (std) | -0.07 | 0.91 | 0.52 | 0.76 | -0.07 | 0.90 | 0.46 | 0.79 | -0.04 | 0.90 | 0.43 | 0.81 |
| COVID-19 stress (std) | 0.08 | 0.92 | 0.59 | 0.71 | 0.01 | 0.86 | 0.55 | 0.71 | -0.23 | 0.72 | 0.46 | 0.71 |
| COVID-19 infection | 0.11 | 0.32 | 0.13 | 0.86 | 0.04 | 0.13 | 0.16 | 0.40 | 0.03 | 0.10 | 0.13 | 0.37 |
| Social support (std) | -0.10 | 0.87 | 0.37 | 0.85 | -0.03 | 0.92 | 0.32 | 0.89 | -0.02 | 0.94 | 0.31 | 0.90 |

Notes: All predictors were standardised (std) in the total sample to have a mean of 0 and standard deviation of 1, except for the binary variable, COVID-19 infection.

### Mental health changes over time

Figure 1 shows the weighted mean of anxiety and depressive symptoms and its 95% confidence interval from each wave in relation to stringency index, average daily COVID-19 cases and deaths (see Figure S2 for categorised measures). Both depressive and anxiety symptoms were high at the start of the first national lockdown, but decreased rapidly in the first few months before starting to raise again in August 2020 until the end of the third national lockdown in March 2021. Both measures then decreased gradually into the summer months of 2021, but then showed increases towards December 2021-January 2022. This is particularly the case for depressive symptoms which lowered again in March-April 2022. These trends were consistent with descriptive statistics presented above, but showing detailed gradients in changes over time.

**Figure 1.**
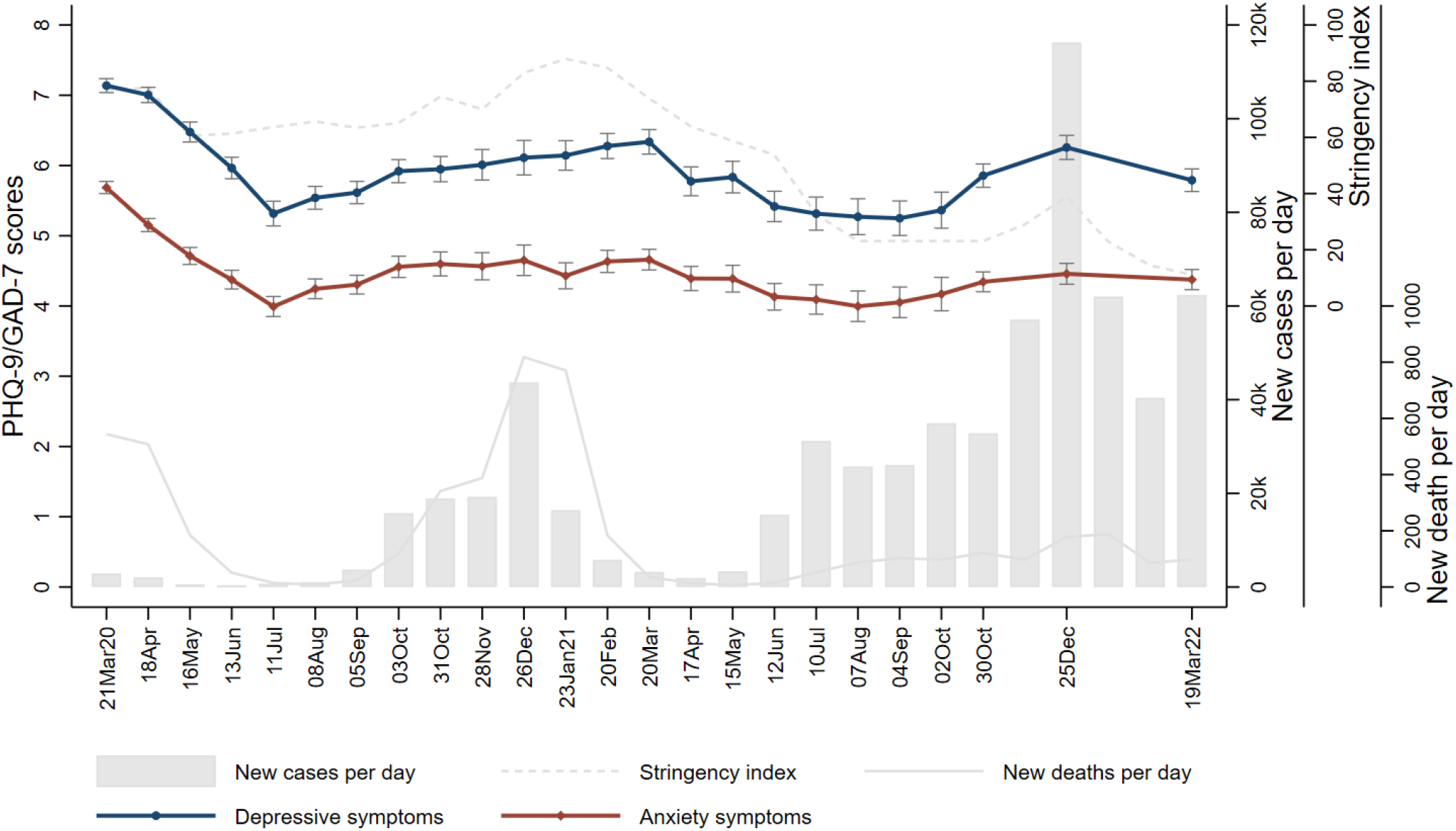
Trends of depressive and anxiety symptoms (weighted means) over time from March 2020 to March 2022

### Longitudinal Predictors of changes in mental health

As expected, most contextual measures were closely related to each other (Figure S3; see Table S2 for multicollinearity diagnostics). Therefore, it is important to examine the longitudinal association between each predictor and the outcomes independent of other factors. The results from FE models including both contextual and individual level factors are reported in Table 2 and Figure 2. Both p and q values are reported in Table 2. We would refer to q values when interpreting the results.

**Table 2.**
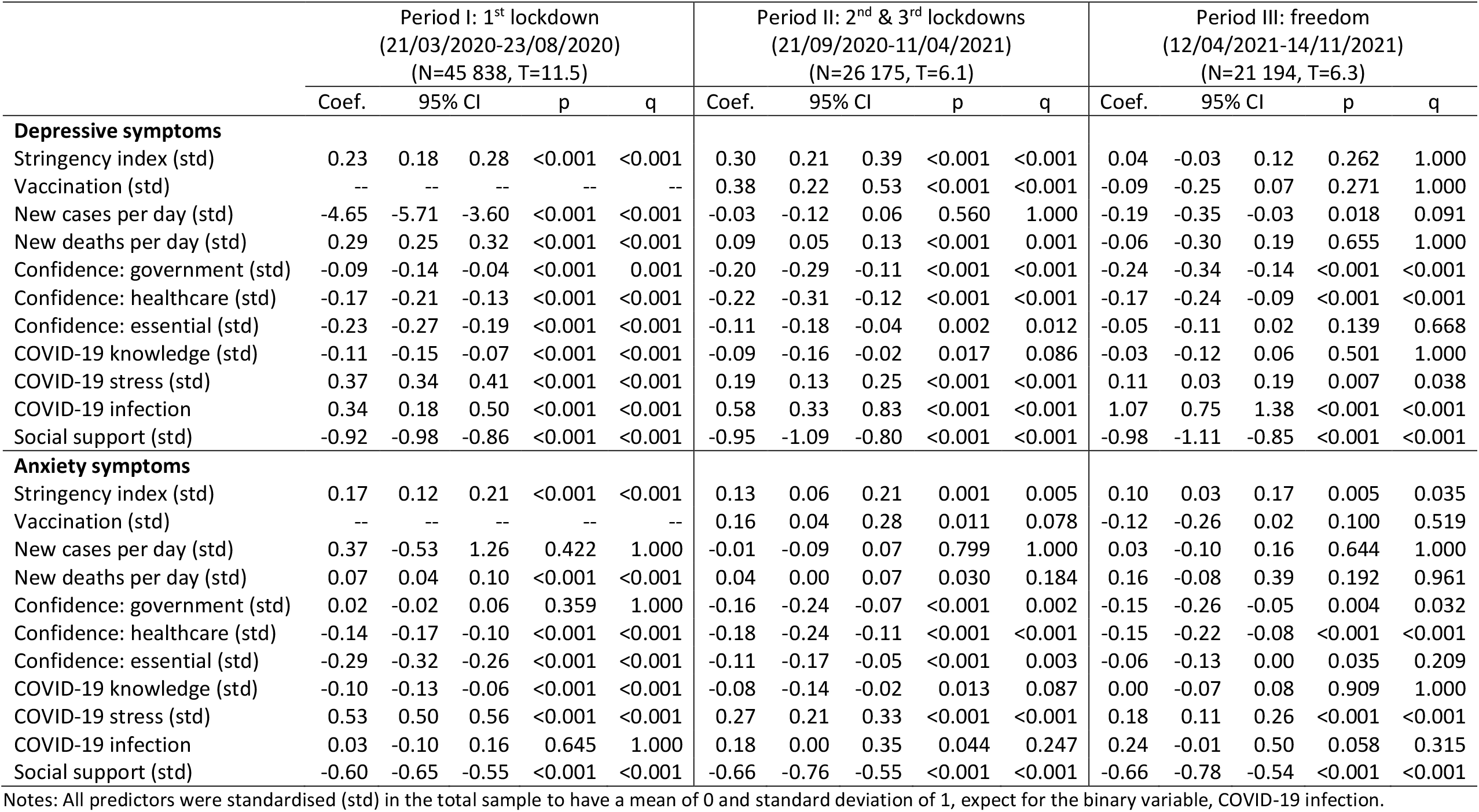
Results from fixed effects models across three periods (weighted)

**Figure 2.**
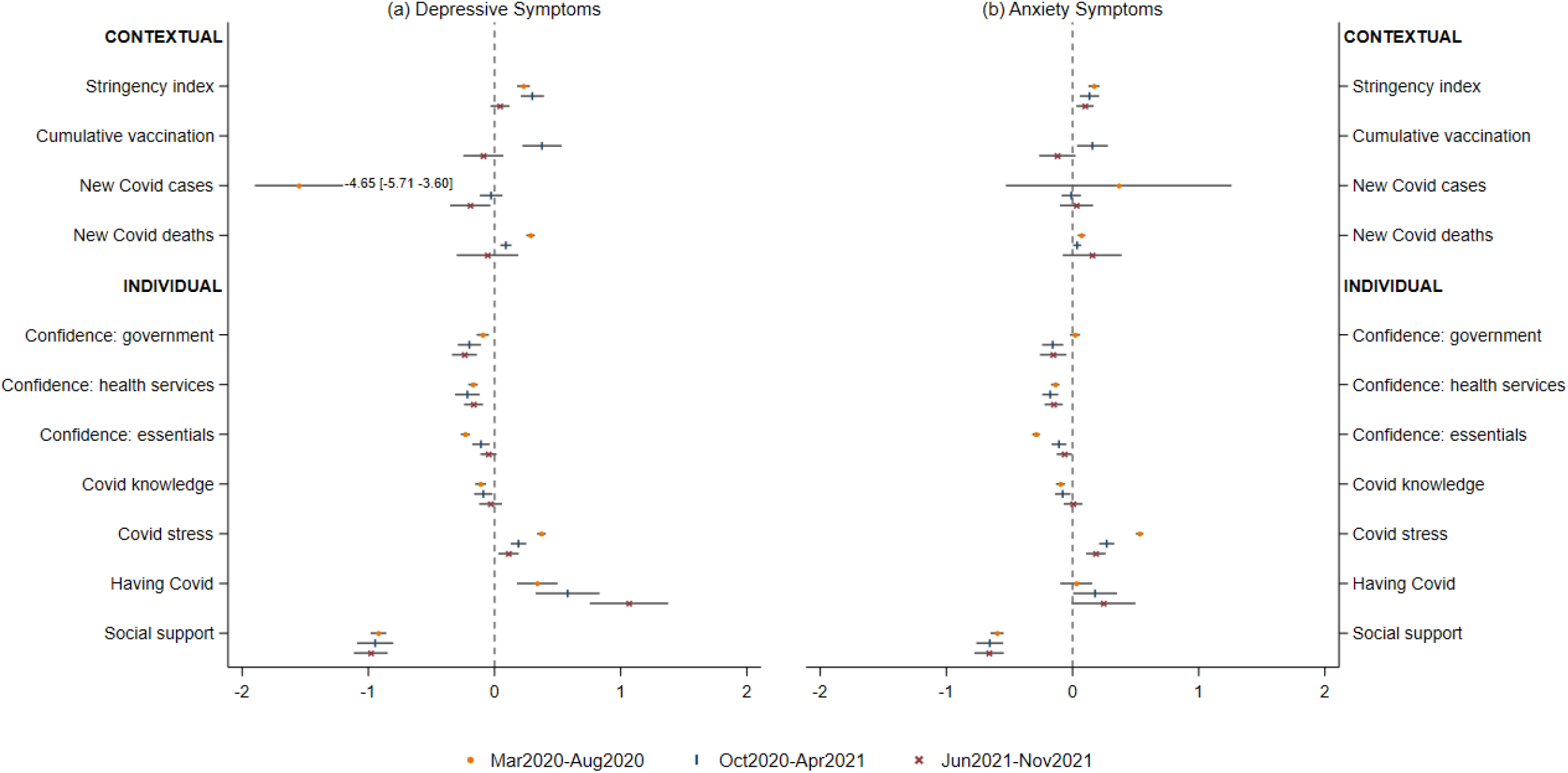
Estimated coefficients and 95% confidence intervals from fixed effects models of three time periods during the COVID-19 pandemic Note: the estimates for new Covid case from models on depressive symptoms were rescaled to fall within the chosen range.

#### Depressive symptoms

During Periods I and II, one standardised deviation (SD) increase in stringency index was associated with 0.23-0.30 points increase in depressive symptoms. But there was no evidence for this longitudinal association in Period III. An increase in COVID-19 cases was associated with a decrease in depressive symptoms, but only in Period I. An increase in COVID-19 deaths was associated with higher depressive symptoms, but this abated over time. One SD increase in vaccination was associated with a 0.38-point increase in depressive symptoms in Period II, without any evidence in Period III.

There was a negative longitudinal association between confidence in government and depressive symptoms across all study periods, with the magnitude of the association increasing over time. A negative longitudinal association with depressive symptoms was also found for confidence in healthcare services in all periods, and for confidence in access to essentials in Periods I and II (although the latter decreased in strength over time). One SD increase in COVID-19 knowledge was associated with a 0.11-point decrease in depressive symptoms in Period I. But no evidence was found in Periods II and III. An increase in COVID-19 related stress was associated with an increase in depressive symptoms, with the association being smaller in later periods. COVID-19 infection was associated with increases in depressive symptoms in all periods, with this association becoming stronger in later periods. Finally, increases in social support were consistently associated with decreases in depressive symptoms.

#### Anxiety symptoms

Increases in stringency index were associated with increases in anxiety symptoms in all study periods. There was no evidence for a longitudinal association of anxiety with COVID-19 cases. However, one SD increase in COVID-19 deaths was associated a 0.07-point increase in anxiety, but in Period I only. There was limited evidence for a longitudinal association between national vaccination and anxiety. There was no evidence that confidence in government was associated with anxiety in Period I, but significant longitudinal associations were found in Periods II and III. A negative longitudinal association of anxiety with confidence in healthcare services was found in all study periods; whereas its relationship with confidence in essentials was significant in Periods I and II only. Increased COVID-19 knowledge was associated with lower anxiety in Period I, with limited evidence in Periods II and III. Increases in COVID-19 stress were associated with higher anxiety symptoms consistently, although the association became less prominent in later periods. There was no evidence for a longitudinal association between COVID-19 infection and anxiety. Finally, increased social support was associated with lower anxiety symptoms across time.

## Discussion

Drawing data from a large panel study with regular follow-ups, this study showed how depressive and anxiety symptoms changed by month over two years between March 2020 and April 2022 in England. This makes it the UK study with the longest and most frequent mental health follow-ups after the outbreak of the COVID-19 pandemic. Both depressive and anxiety symptoms were high at the start of the first national lockdown, but fell rapidly in the first few months. However, they rose in late 2020 to early 2021 as the number of cases increased and COVID-19 restrictions tightened before deceasing again following the end of the third national lockdown, despite relatively high numbers of cases. This is generally in line with findings from other data [12]. In addition, our study showed that there was a rise in depression in particular in late 2021 when case numbers peaked, even though no further lockdowns were brought in. Further, our analyses showed how changes in contextual and individual level factors were associated with changes in depressive and anxiety symptoms across different stages of the pandemic, including the first national lockdown, the second and third national lockdowns, and the period of minimal restrictions.

Several factors were related to both depressive and anxiety symptoms consistently across all study periods. These included confidence in healthcare services, COVID-19 related stress, and perceived social support. In the face of the COVID-19 crisis, the National Health Service (NHS) has been under extreme pressure because of surging demands and staff absences [22]. This was reinforced by the fact that ‘protect the NHS’ was communicated as part of the key public messages at the COVID-19 press briefings at the early stage of the pandemic. Therefore, it is not surprising that people’s confidence in how the NHS coped during the pandemic was related to depressive and anxiety symptoms, regardless of time periods. Additionally, there was evidence to suggest that during the pandemic, many people experienced disruptions to accessing healthcare services or reductions in help-seeking due to fear of COVID-19 infection or feeling that they should not burden the health service [23, 24]. So perceived unavailability of mental health support due to overall load on the health service could also explain the relationship with higher anxiety and depressive symptoms. It is notable that the relationship between stress about COVID-19 and mental health symptoms weakened over time (even though it remained significant and, compared to other standardised predictors, meaningful in size). This was likely influenced by the roll-out of vaccinations, which reduced the risk of hospitalisation and deaths from the virus. However, the associations with stress about COVID-19 were independent of the vaccination variable. Therefore, the diminishing relationship between COVID-19 stress and mental health over time is likely at least in part due to habituation, whereby the ongoing exposure to information on COVID-19 and increased number of contacts who had contracted and survived COVID-19 reduced the psychological salience of the stressor [25]. Our findings on perceived social support are in line with pre-pandemic literature showing the mental health benefits of social relationships [26, 27]. They suggest that social support continued to play an important role in supporting people’s mental health during the heightened emotional distress and psychological challenges of the COVID-19 pandemic. In fact, social support was arguably the most important predictor overall when comparing coefficients across standardised predictors.

Other factors showed differential longitudinal association with depressive and/or anxiety symptoms across different stages of the pandemic. For instance, increases in COVID-19 policy measures were associated with increased depressive symptoms during lockdown periods, but no evidence was found after the easing of restrictions following the third national lockdown. This could be because policy changes at this stage were relatively minor, which had limited impact on personal freedom especially after most legal restrictions being removed in July 2021. It also suggests that the relationship between policy stringency and mental health may not be proportionate, with increases in policy changes becoming more damaging for mental health as they infringe more on social freedoms. Increased COVID-19 deaths were associated with higher depressive and anxiety symptoms only at early stages of the pandemic. This might be explained by the relatively low and stable death rate in the ‘freedom’ period possibly due to the roll out of vaccination programme, so that deaths from COVID-19 no longer presented itself as a major psychological threat. However, after controlling for other contextual factors, our analyses showed that progresses in vaccination programme was associated with higher depressive symptoms and anxiety (non-significant q value) in the second study period. The vaccination programme in England began on 8 December 2020. There had been significant vaccine scepticism and hesitancy at the start [22, 28], which may explain the positive longitudinal association. However, with effective communication of scientific information on COVID-19 vaccination over time, this association was no longer observed at the later stage.

For individual level factors, increased confidence in accessing essentials was associated with reduced depressive and anxiety symptoms only during lockdown periods, particularly the first national lockdown when essential and non-essential services were most heavily influenced either due to COVID-19 restrictions or staff shortage. Finally, an increase in self-rated COVID-19 knowledge was associated with decreased depressive and anxiety symptoms, but only in the first lockdown period. A plausible explanation is that knowledge about the virus and its transmission was particularly salient at the start when there was a general lack of understanding accompanied by excessive circulation of misinformation [29].

Although depressive and anxiety symptoms are highly correlated with each other, it is important to acknowledge differences in how they are related to some of the predictors. Our findings showed that COVID-19 infection was associated with increased depressive symptoms consistently across all study periods, but limited evidence was found for anxiety. Notably, this association was independent of stress about COVID-19. Further, it is interesting that that association was for having COVID-19 oneself rather than the overall national case rates of COVID-19. This suggests that individual psychobiological mechanisms of COVID-19 could be responsible for the association with depressive symptoms. Existing literature proposes that inflammatory mechanisms in COVID-19 infection could be responsible for an increase in depressive symptoms [30, 31]. This could certainly resonate with the association only being found for depression, and not for anxiety [32]. Although there was no evidence for the longitudinal association of stringency index with depressive symptoms in the ‘freedom’ period, the association was found for anxiety symptoms. It is possible that easing of restrictions at this stage despite being inconsequential for depressive symptoms still had some impact on anxiety, relating to COVID-19 or other challenges. Moreover, confidence in government was associated with depressive symptoms in all periods, but no evidence was found during the first national lockdown period for anxiety. Relatively speaking, there was a higher level of support for COVID-19 policy responses at the start of the pandemic as shown in confidence in government[33]; whereas people disagreed to a greater extent with the government at later stages (Table 1). Therefore, it is possible that people were less anxious in relation to confidence in government (after controlling for policy stringency) during the first national lockdown when confidence in their capability was higher.

This study has a number of strengths. It utilised a large sample with sufficient heterogeneity to include good stratification across all major socio-demographic groups and good coverage of geographic areas in England. The analyses were weighted on the basis of population estimates of core demographics, with the weighted data showing good alignment with a nationally representative study. Due to the longitudinal design of the COVID-19 Social Study, we were able to examine overall trends of depressive and anxiety symptoms over 25 months, and longitudinal predictors of changes in them across different stages of the COVID-19 pandemic. However, our study is not without any limitation. First, it is important to acknowledge that our data were from a non-probability sample. Despite the effort to recruit a heterogeneous sample and make our sample representative to the adult population in England by weighting, there is still the possibility of potential biases due to omitting other demographic factors that could be associated with survey participation in the weighting process. We therefore advice caution when generalising these findings to the population. Moreover, the fixed effects approach cannot rule out the possibility of bidirectional associations of depressive/anxiety symptoms with individual level factors. This calls for further research in this area.

Since July 2021, England removed most of its legal restrictions, making it one of the countries with the least strict COVID-19 policies (see Figure S4) and highest vaccine coverage (Figure S5) at the time of writing. The United Kingdom is also one of the countries with highest cumulative number of COVID-19 cases and deaths (Figure S6-S7). But there is a lot of variation in COVID-19 situation and policy stringency across countries and over time. Our results provided empirical evidence on how changes in contextual measures, including stringency index, COVID-19 cases, COVID-19 deaths and national vaccination, as well as individual level factors, such as COVID-19 related stress, COVID-19 infection and social support were related to depressive and anxiety symptoms. While some factors (especially individual factors) clearly act as consistent predictors of mental health during a pandemic (e.g. social support, stress about catching the virus, and confidence in government, health services and access to essentials), other factors (especially contextual factors) are dependent on the specific situations occurring within society as to whether they impact mental health or not (e.g. policy stringency, cases, deaths and vaccination rates). This could provide important implications for policy making and for a better understanding of mental health of the general public during a national or global health crisis.

## Data Availability

Anonymous data will be made publicly available following the end of the pandemic.

## Declarations

### Ethics approval and consent to participate

The study was approved by the UCL Research Ethics Committee [12467/005] and all participants gave informed consent.

### Availability of data and materials

Anonymous data will be made publicly available following the end of the pandemic.

### Author contributions

AS and DF developed the study idea. FB developed the analysis plan and analysed the data. FB and DF wrote the first draft. FB and DF had accessed and verified the underlying data. All authors provided critical revisions, read and approved the submitted manuscript.

## Acknowledgements

The researchers are grateful for the support of a number of organisations with their recruitment efforts including: the UKRI Mental Health Networks, Find Out Now, UCL BioResource, SEO Works, FieldworkHub, and Optimal Workshop. The study was also supported by HealthWise Wales, the Health and Care Research Wales initiative, which is led by Cardiff University in collaboration with SAIL, Swansea University.

## Reference

[1] Carr MJ, Steeg S, Webb RT, et al. Effects of the COVID-19 pandemic on primary care-recorded mental illness and self-harm episodes in the UK: a population-based cohort study. The Lancet Public Health 2021; 6: e124–e135.

[2] Fancourt D, Steptoe A, Bu F. Trajectories of anxiety and depressive symptoms during enforced isolation due to COVID-19 in England: a longitudinal observational study. The Lancet Psychiatry 2021; 8: 141–149.

[3] Holmes EA, O’Connor RC, Perry VH, et al. Multidisciplinary research priorities for the COVID-19 pandemic: a call for action for mental health science. The Lancet Psychiatry 2020; 7: 547–560.

[4] Mahase E. Covid-19: Mental health consequences of pandemic need urgent research, paper advises. BMJ 2020; 369: m1515.

[5] Pierce M, Hope H, Ford T, et al. Mental health before and during the COVID-19 pandemic: a longitudinal probability sample survey of the UK population. The Lancet Psychiatry 2020; 7: 883–892.

[6] Daly M, Sutin AR, Robinson E. Longitudinal changes in mental health and the COVID-19 pandemic: evidence from the UK Household Longitudinal Study. Psychological Medicine 2020; 1–10.

[7] Kwong ASF, Pearson RM, Adams MJ, et al. Mental health before and during the COVID-19 pandemic in two longitudinal UK population cohorts. Br J Psychiatry 2020; 1–10.

[8] Robinson E, Sutin AR, Daly M, et al. A systematic review and meta-analysis of longitudinal cohort studies comparing mental health before versus during the COVID-19 pandemic in 2020. Journal of Affective Disorders 2022; 296: 567–576.

[9] Pierce M, McManus S, Hope H, et al. Mental health responses to the COVID-19 pandemic: a latent class trajectory analysis using longitudinal UK data. The Lancet Psychiatry 2021; 8: 610–619.

[10] Lee Y, Lui LMW, Chen-Li D, et al. Government response moderates the mental health impact of COVID-19: A systematic review and meta-analysis of depression outcomes across countries. Journal of Affective Disorders 2021; 290: 364–377.

[11] de la Rosa PA, Cowden RG, de Filippis R, et al. Associations of lockdown stringency and duration with Google searches for mental health terms during the COVID-19 pandemic: A nine-country study. Journal of Psychiatric Research 2022; 150: 237–245.

[12] Aknin LB, Andretti B, Goldszmidt R, et al. Policy stringency and mental health during the COVID-19 pandemic: a longitudinal analysis of data from 15 countries. The Lancet Public Health 2022; 7: e417–e426.

[13] Wright L, Fancourt D. Do predictors of adherence to pandemic guidelines change over time? A panel study of 22,000 UK adults during the COVID-19 pandemic. Preventive Medicine 2021; 153: 106713.

[14] Löwe B, Kroenke K, Herzog W, et al. Measuring depression outcome with a brief self-report instrument: sensitivity to change of the Patient Health Questionnaire (PHQ-9). Journal of Affective Disorders 2004; 81: 61–66.

[15] Spitzer RL, Kroenke K, Williams JBW, et al. A Brief Measure for Assessing Generalized Anxiety Disorder. Archives of Internal Medicine 2006; 166: 1092.

[16] Hale T, Angrist N, Goldszmidt R, et al. A global panel database of pandemic policies (Oxford COVID-19 Government Response Tracker). Nature Human Behaviour 2021 5:4 2021; 5: 529–538.

[17] UK Health Security Agency. Coronavirus (COVID-19) in the UK, https://coronavirus.data.gov.uk/ (2022, accessed May 23, 2022).

[18] Kliem S, Mößle T, Rehbein F, et al. A brief form of the Perceived Social Support Questionnaire (F-SozU) was developed, validated, and standardized. Journal of Clinical Epidemiology 2015; 68: 551–562.

[19] Office for National Statistics. Population estimates for the UK, England and Wales, Scotland and Northern Ireland, https://www.ons.gov.uk/peoplepopulationandcommunity/populationandmigration/populationestimates/bulletins/annualmidyearpopulationestimates/mid2018 (2019, accessed May 13, 2020).

[20] Wooldridge JM. Econometric Analysis of Cross Section and Panel Data. 2nd ed. MIT Press, 2010.

[21] Newson RB. Frequentist Q-values for Multiple-test Procedures: https://doi.org/101177/1536867X1101000403 2011; 10: 568–584.

[22] Awijen H, ben Zaied Y, Nguyen DK. Covid-19 vaccination, fear and anxiety: Evidence from Google search trends. Social Science & Medicine 2022; 297: 114820.

[23] World Health Organization. The impact of COVID-19 on mental, neurological and substance use services: results of a rapid assessment. 2020.

[24] Chen S, Jones PB, Underwood BR, et al. The early impact of COVID-19 on mental health and community physical health services and their patients’ mortality in Cambridgeshire and Peterborough, UK. Journal of Psychiatric Research 2020; 131: 244–254.

[25] Eisenstein EM, Eisenstein D. A behavioral homeostasis theory of habituation and sensitization: II. Further developments and predictions. Reviews in the Neurosciences 2006; 17: 533–557.

[26] Uchino BN, Cacioppo JT, Kiecolt-Glaser JK. The relationship between social support and physiological processes: a review with emphasis on underlying mechanisms and implications for health. Psychol Bull 1996; 119: 488–531.

[27] Wang J, Mann F, Lloyd-Evans B, et al. Associations between loneliness and perceived social support and outcomes of mental health problems: A systematic review. BMC Psychiatry 2018; 18: 1–16.

[28] Paul E, Steptoe A, Fancourt D. Attitudes towards vaccines and intention to vaccinate against COVID-19: Implications for public health communications. The Lancet Regional Health - Europe 2021; 1: 100012.

[29] Al-Zaman MS. Prevalence and source analysis of COVID-19 misinformation in 138 countries: https://doi.org/101177/03400352211041135 2021; 48: 189–204.

[30] Jokela M, Virtanen M, David Batty G, et al. Inflammation and Specific Symptoms of Depression. JAMA Psychiatry 2016; 73: 87–88.

[31] da Silva Lopes L, Silva RO, de Sousa Lima G, et al. Is there a common pathophysiological mechanism between COVID-19 and depression? Acta Neurologica Belgica 2021; 121: 1117–1122.

[32] Fancourt D, Steptoe A, Bu F. Long-term psychological consequences of long Covid: a propensity score matching analysis comparing trajectories of depression and anxiety symptoms before and after contracting long Covid vs short Covid. medRxiv 2022; 2022.04.01.22273305.

[33] Seyd B, Bu F. Perceived risk crowds out trust? Trust and public compliance with coronavirus restrictions over the course of the pandemic. European Political Science Review 2022; 1–16.

