## Supplement for "Depressive and anxiety symptoms during the COVID-19 pandemic: A two-year follow-up"

Table S1 Baseline sample characteristics across three study periods (weighted)

| | Period I: 1st lockdown (21/03/2020-23/08/2020) (N=45 838) | | Period II: 2nd & 3rd lockdowns (21/09/2020-11/04/2021) (N=26 175) | | Period III: freedom (12/04/2021-14/11/2021) (N=21 194) | |
|---|---|---|---|---|---|---|
| | Percentage | Frequency | Percentage | Frequency | Percentage | Frequency |
| **Gender** | | | | | | |
| Women | 50.2% | 23289 | 50.5% | 13317 | 50.1% | 10774 |
| Men | 49.8% | 22549 | 49.5% | 12858 | 49.9% | 10420 |
| **Age** | | | | | | |
| 18-29 | 18.4% | 8983 | 17.6% | 5124 | 17.5% | 4151 |
| 30-45 | 27.7% | 12101 | 27.7% | 6903 | 27.9% | 5593 |
| 46-59 | 24.4% | 11003 | 24.7% | 6290 | 24.7% | 5089 |
| 60+ | 29.6% | 13750 | 30.1% | 7857 | 29.9% | 6361 |
| **Ethnicity** | | | | | | |
| Ethnic minority groups | 14.2% | 6692 | 14.0% | 3817 | 14.1% | 3092 |
| White | 85.8% | 39146 | 86.0% | 22358 | 85.9% | 18102 |
| **Education** | | | | | | |
| Low (GCSEs or below) | 32.1% | 14985 | 31.9% | 8536 | 31.8% | 6922 |
| Medium (A-levels or equivalent) | 32.6% | 15168 | 31.9% | 8644 | 32.1% | 7007 |
| High (Degree or above) | 35.3% | 15685 | 36.1% | 8995 | 36.1% | 7265 |
| **Household income** | | | | | | |
| <30k | 45.9% | 18819 | 46.2% | 10647 | 46.9% | 8735 |
| ≥30k | 54.1% | 22190 | 53.8% | 12393 | 53.1% | 9895 |
| **Employed status** | | | | | | |
| Employed | 60.2% | 26591 | 59.1% | 14845 | 59.8% | 12067 |
| Other | 39.8% | 19247 | 40.9% | 11330 | 40.2% | 9127 |
| **Mental health diagnosis** | | | | | | |
| Yes | 20.1% | 9155 | 19.2% | 4986 | 19.0% | 3987 |
| No | 79.9% | 36683 | 80.8% | 21189 | 81.0% | 17207 |

Table S2 Multicollinearity diagnostics across study periods (unweighted)

| | Period I (n=526818) | | Period II (n=160250) | | Period III (n=133892) | |
|---|---|---|---|---|---|---|
| | VIF | Tolerance | VIF | Tolerance | VIF | Tolerance |
| Stringency index (std) | 2.95 | 0.34 | 2.77 | 0.36 | 6.11 | 0.16 |
| Vaccination (std) | -- | -- | 2.40 | 0.42 | 7.06 | 0.14 |
| New cases per day (std) | 5.86 | 0.17 | 2.03 | 0.49 | 4.07 | 0.25 |
| New deaths per day (std) | 4.16 | 0.24 | 2.62 | 0.38 | 2.13 | 0.47 |
| Confidence: government (std) | 1.41 | 0.71 | 1.28 | 0.78 | 1.30 | 0.77 |
| Confidence: healthcare (std) | 1.80 | 0.56 | 1.84 | 0.54 | 1.83 | 0.55 |
| Confidence: essential (std) | 1.69 | 0.59 | 1.81 | 0.55 | 2.02 | 0.50 |
| COVID knowledge (std) | 1.06 | 0.95 | 1.04 | 0.96 | 1.05 | 0.95 |
| COVID stress (std) | 1.07 | 0.93 | 1.08 | 0.93 | 1.06 | 0.94 |
| COVID infection | 1.01 | 0.99 | 1.00 | 1.00 | 1.00 | 1.00 |
| Social support (std) | 1.06 | 0.94 | 1.06 | 0.94 | 1.06 | 0.95 |

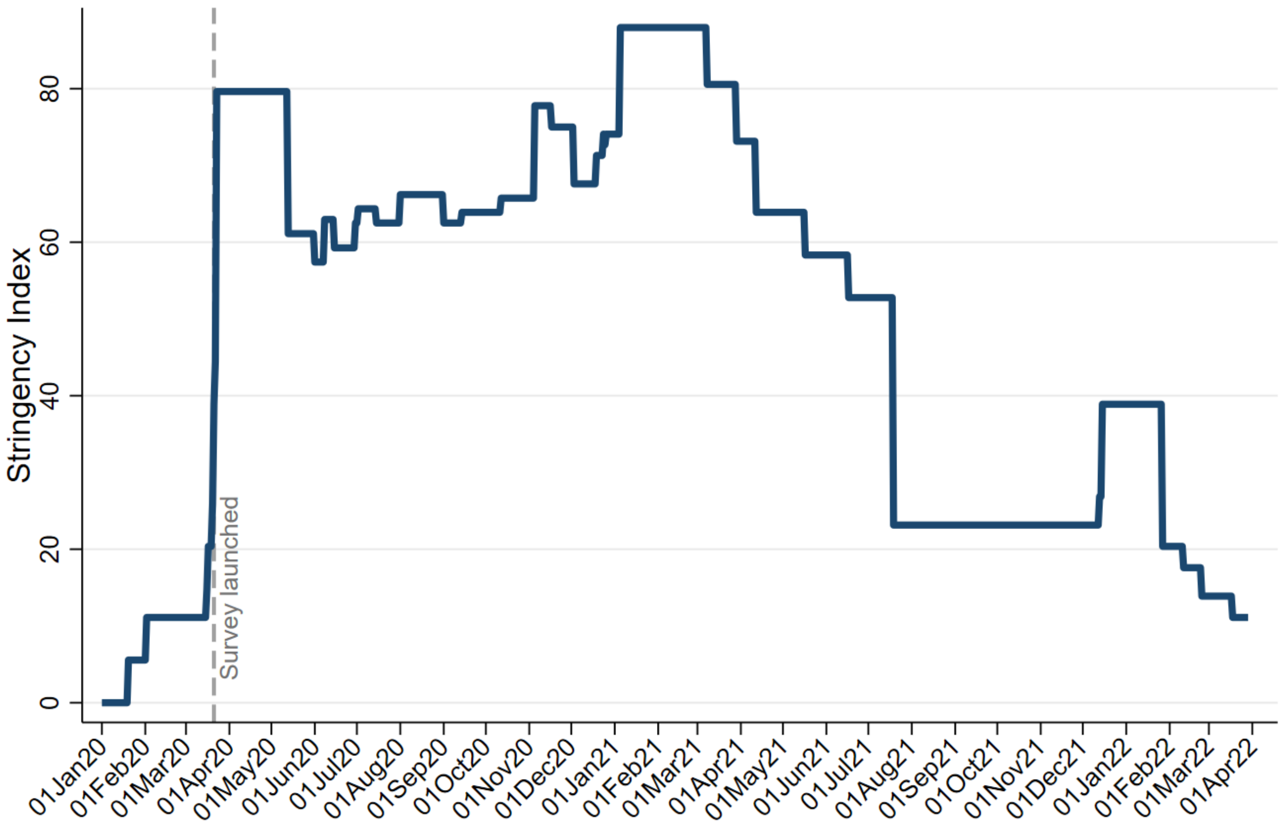


Figure S1 Daily changes in COVID-19 policy responses (stringency index) in England from January 2020 to April 2022

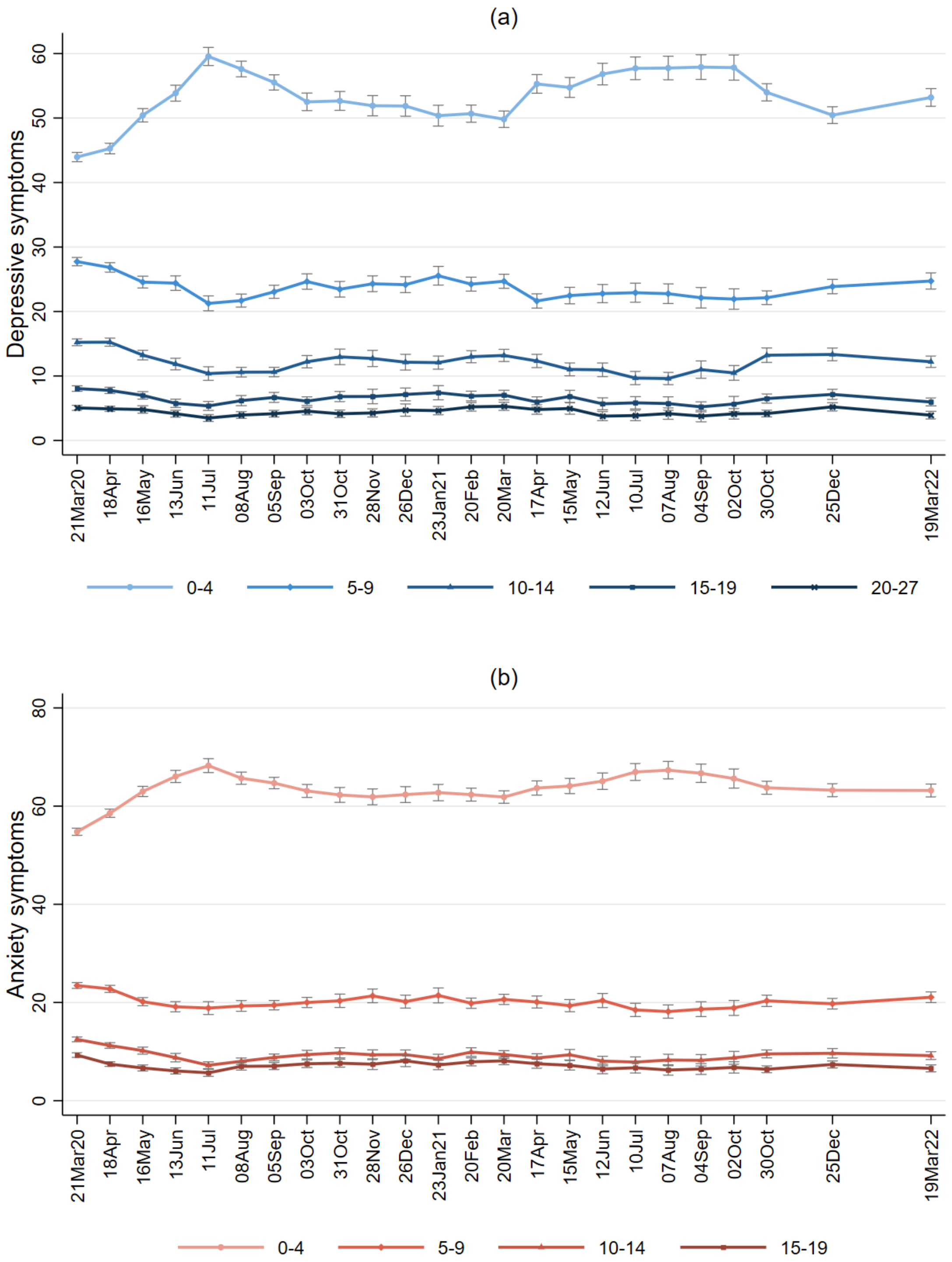


Figure S2 Trends of depressive and anxiety symptoms (weighted percentages in categories) over time from March 2020 to March 2022

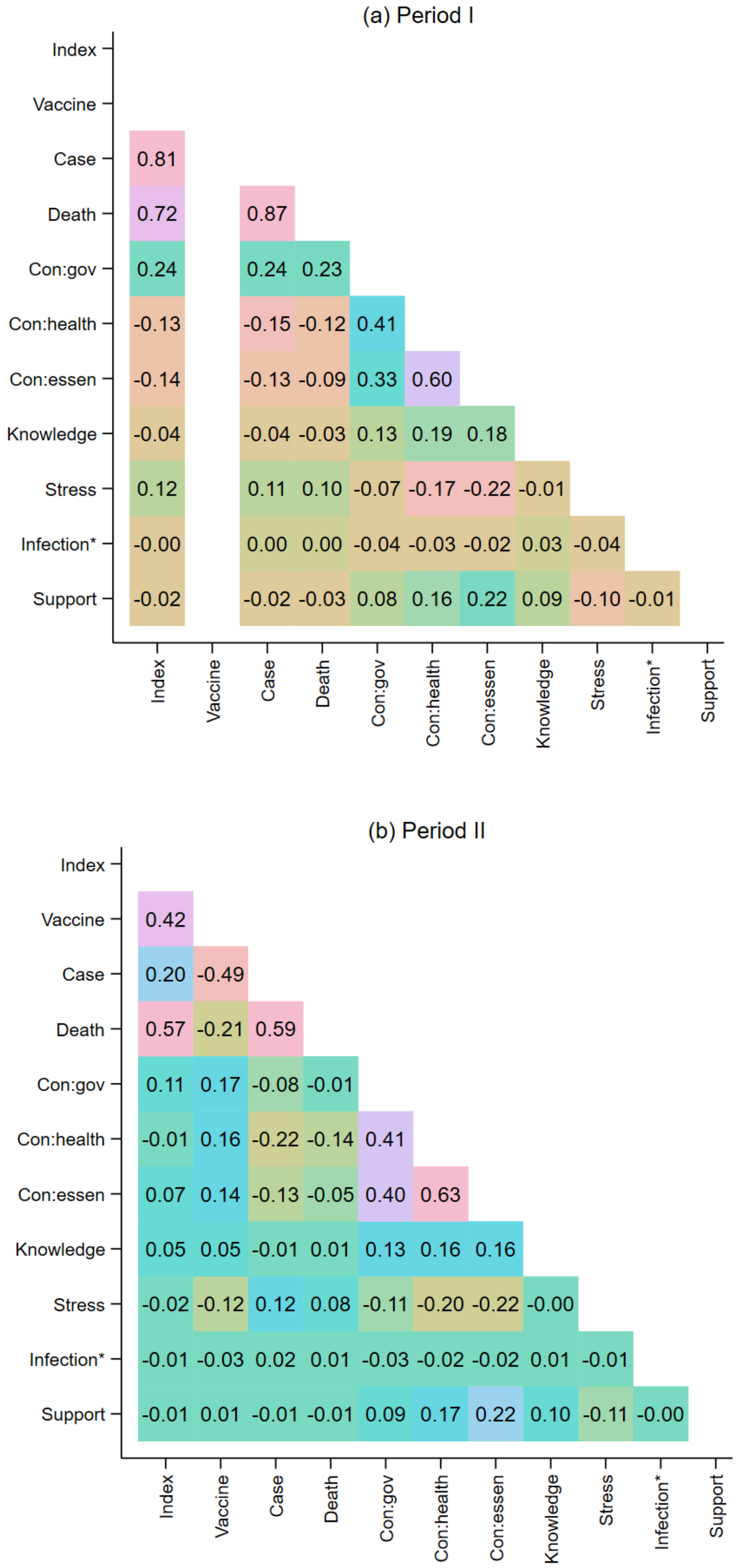
(a) Period I
Index
Vaccine
Case 0.81
Death 0.72 0.87
Con:gov 0.24 0.24 0.23
Con:health -0.13 -0.15 -0.12 0.41
Con:essen -0.14 -0.13 -0.09 0.33 0.60
Knowledge -0.04 -0.04 -0.03 0.13 0.19 0.18
Stress 0.12 0.11 0.10 -0.07 -0.17 -0.22 -0.01
Infection* -0.00 0.00 0.00 -0.04 -0.03 -0.02 0.03 -0.04
Support -0.02 -0.02 -0.03 0.08 0.16 0.22 0.09 -0.10 -0.01
Index Vaccine Case Death Con:gov Con:health Con:essen Knowledge Stress Infection* Support
(b) Period II
Index
Vaccine 0.42
Case 0.20 -0.49
Death 0.57 -0.21 0.59
Con:gov 0.11 0.17 -0.08 -0.01
Con:health -0.01 0.16 -0.22 -0.14 0.41
Con:essen 0.07 0.14 -0.13 -0.05 0.40 0.63
Knowledge 0.05 0.05 -0.01 0.01 0.13 0.16 0.16
Stress -0.02 -0.12 0.12 0.08 -0.11 -0.20 -0.22 -0.00
Infection* -0.01 -0.03 0.02 0.01 -0.03 -0.02 -0.02 0.01 -0.01
Support -0.01 0.01 -0.01 -0.01 0.09 0.17 0.22 0.10 -0.11 -0.00
Index Vaccine Case Death Con:gov Con:health Con:essen Knowledge Stress Infection* Support

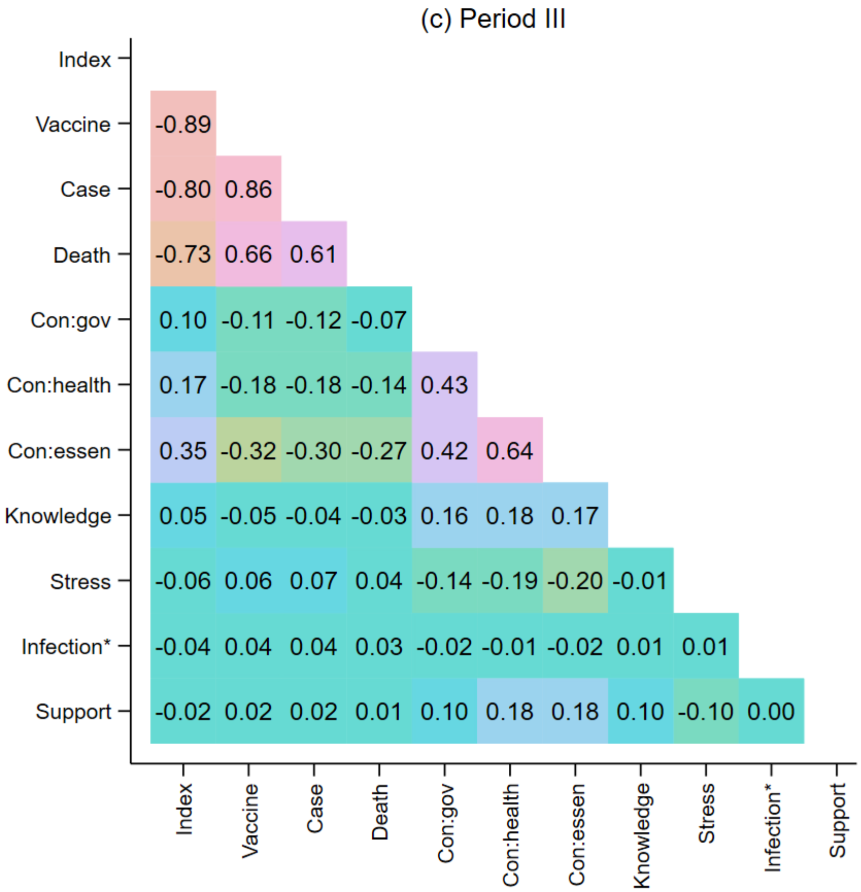


Figure S3 Correlation matrix of predictors across study periods (unweighted)

Note: * binary variable

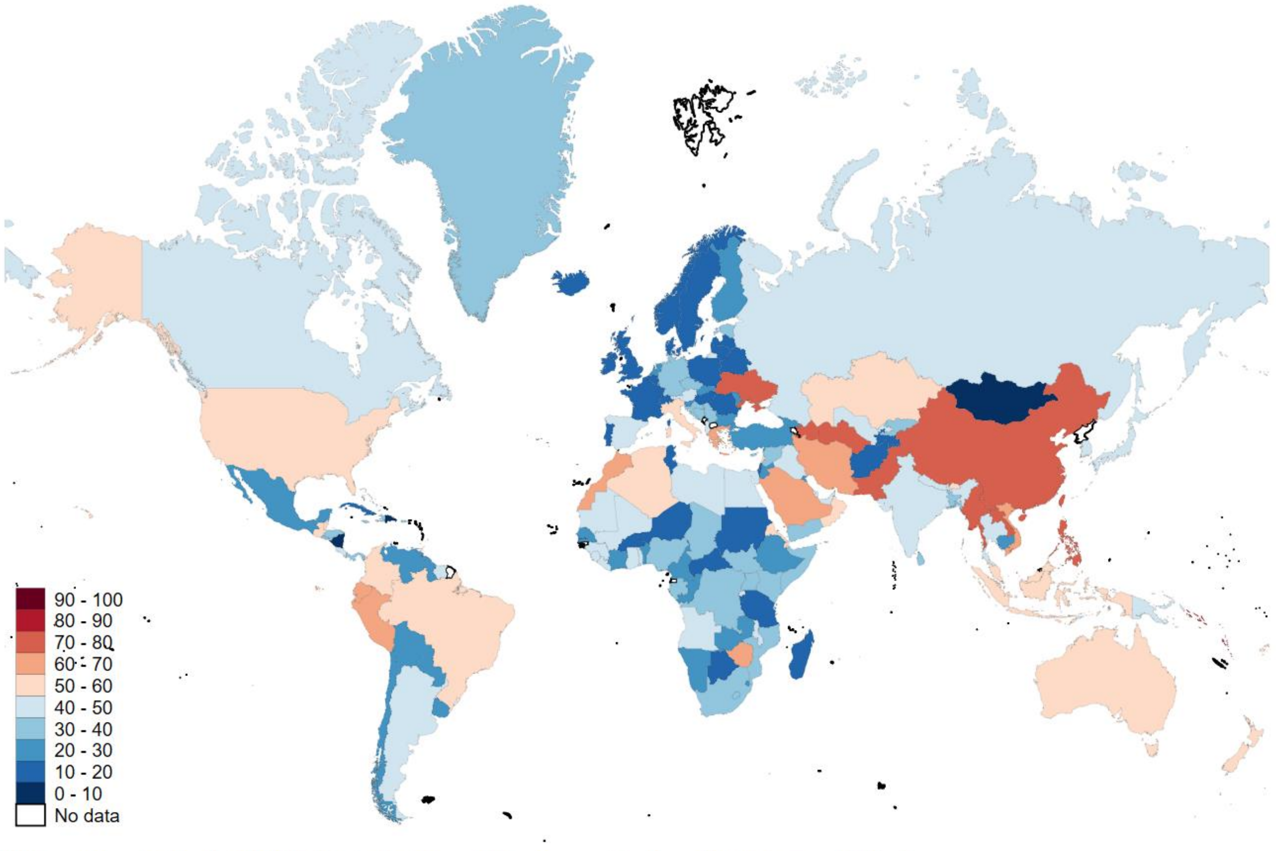


Data source: Oxford COVID-19 Government Response Tracker. Antartica excluded.

Figure S4 Global COVID-19 Policy Stringency Index (20 April 2022)

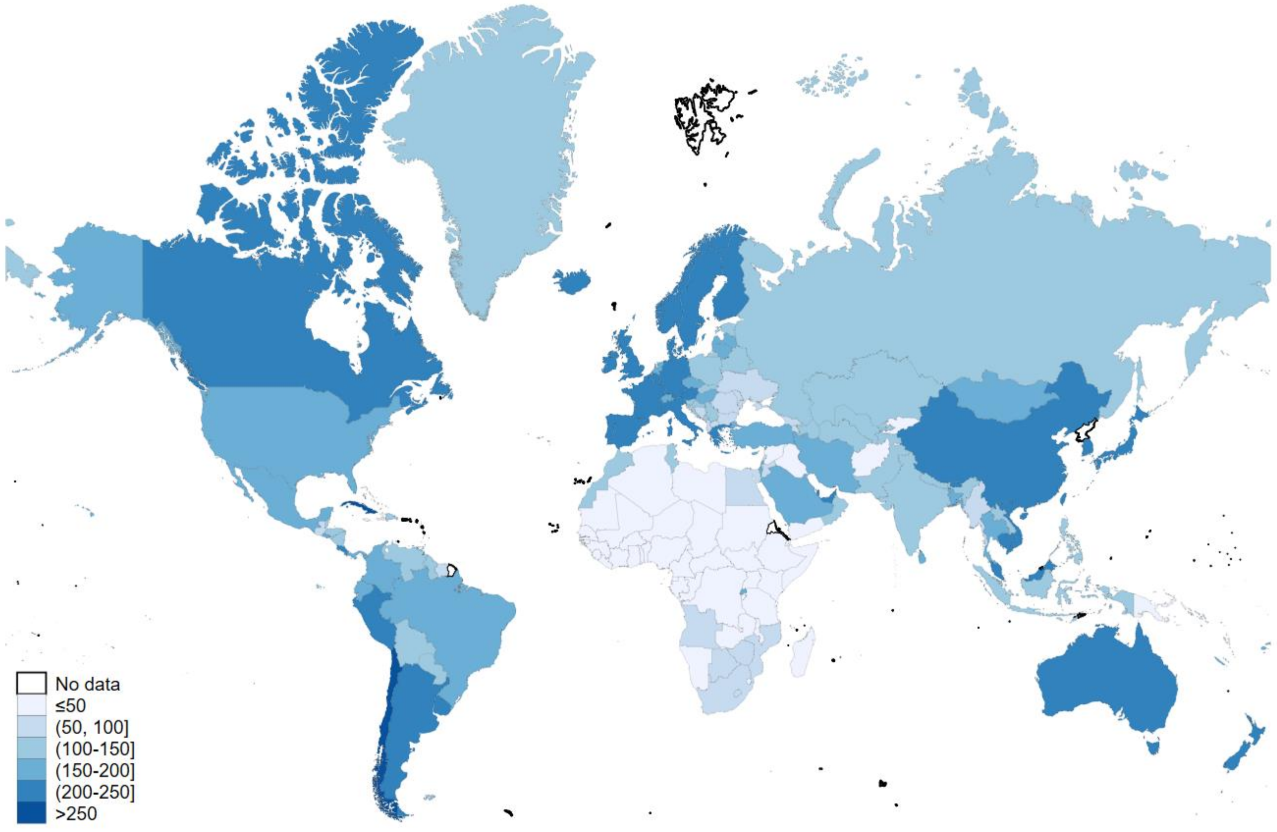


Data source: Our World in Data. Antartica excluded.

Figure S5 Global COVID-19 vaccine does per 100 people (20 April 2022)

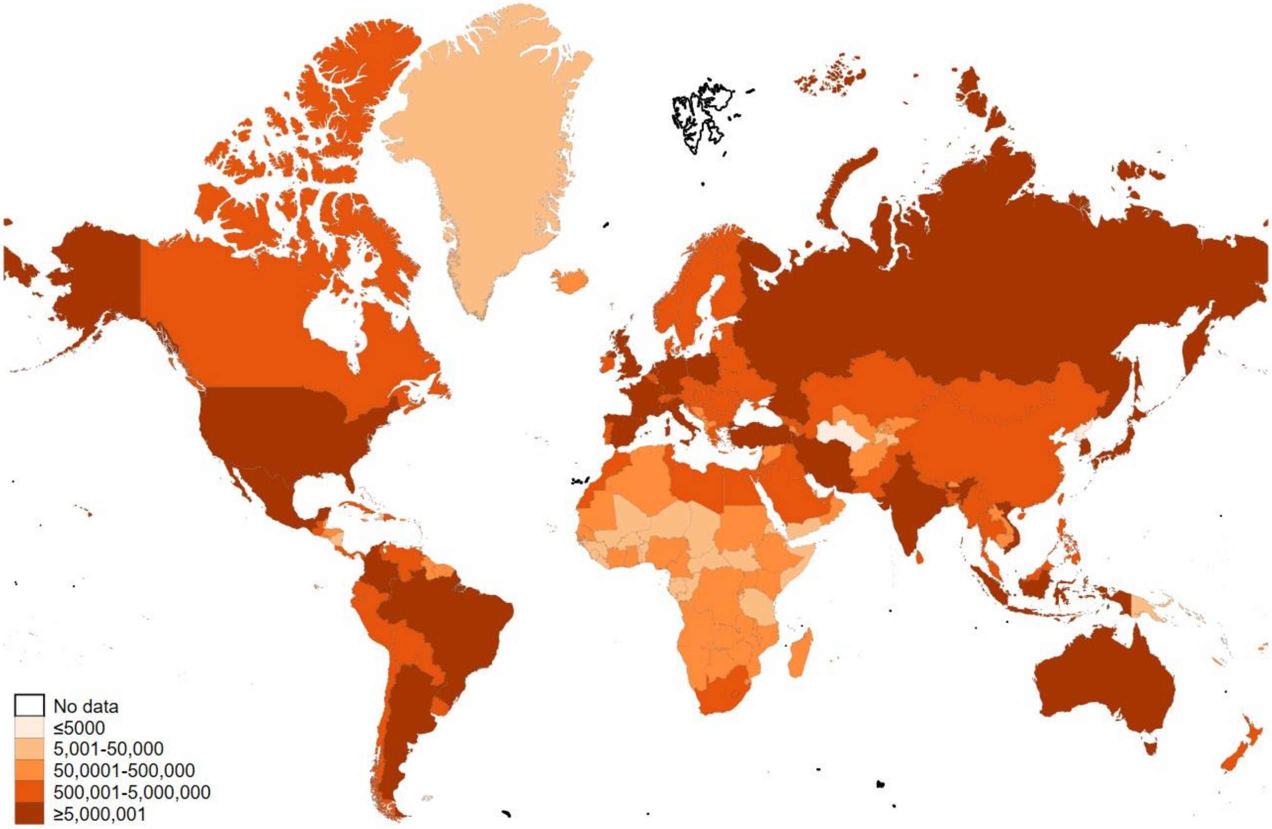


Data source: WHO COVID-19 Dashboard. Antartica excluded.

Figure S6 Global COVID-19 cumulative confirmed cases (19 April 2022)

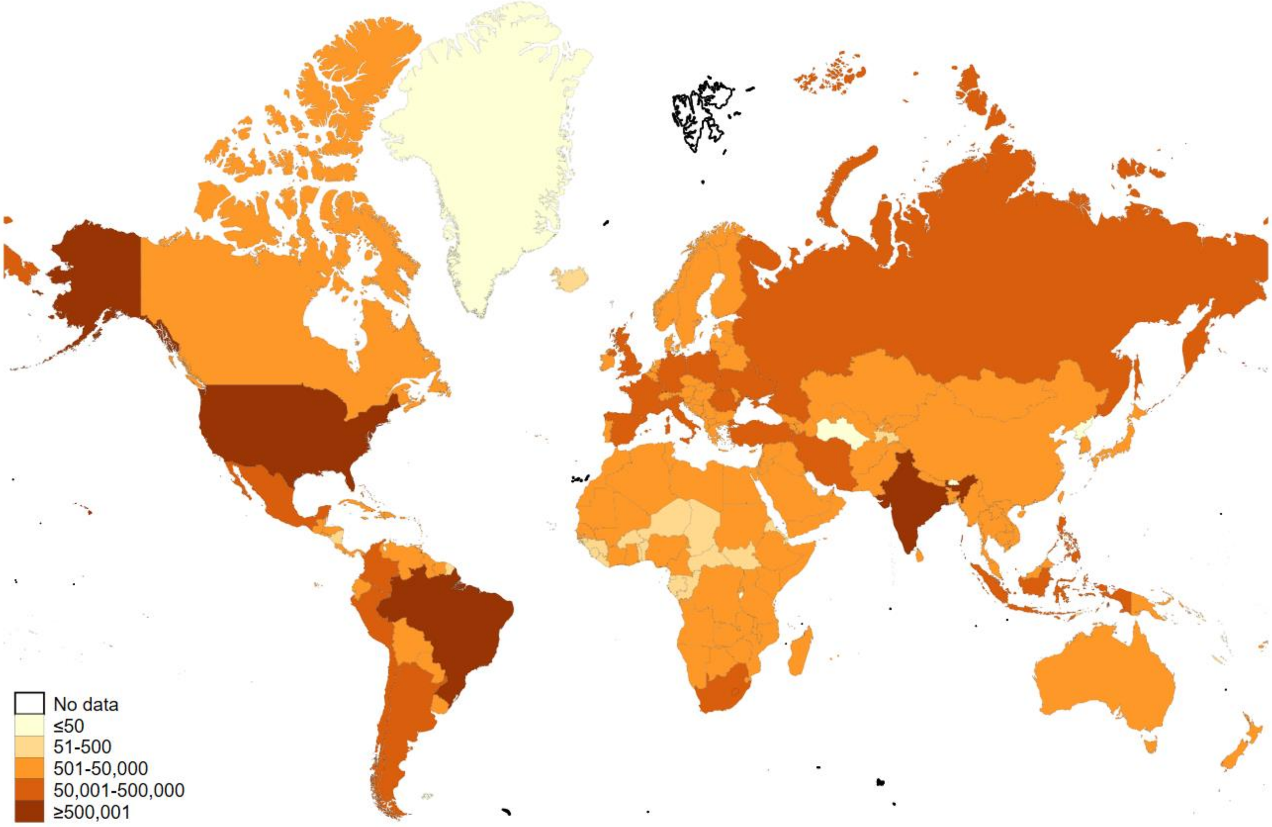


Data source: WHO COVID-19 Dashboard. Antartica excluded.

Figure S7 Global COVID-19 cumulative deaths (19 April 2022)
